# Moving-average based index to timely evaluate the current epidemic situation after COVID-19 outbreak

**DOI:** 10.1101/2020.03.24.20027730

**Authors:** He Yun-ting, Wang Xiao-jin, He Hao, Zhai Jing, Wang Bing-shun

## Abstract

A pneumonia outbreak caused by a novel coronavirus (COVID-19) occurred in Wuhan, China at the end of 2019 and then spread rapidly to the whole country. A total of 81,498 laboratory-confirmed cases, including 3,267 deaths (4.0%) had been reported in China by March 22, 2020, meanwhile, 210,644 laboratory-confirmed cases and 9,517 deaths (4.5%) were reported outside China. Common symptoms of COVID-19 pneumonia included fever, fatigue and dry cough. In face of such a sudden outbreak of emerging novel infectious diseases, we have no history to learn from and no evidence to count on. Traditional models often predict inconsistent results. There is an urgent need to establish a practical data-driven method to predict the evolutionary trend of the epidemic, track and prejudge the current epidemic situation after the COVID-19 outbreak. Here we propose a simple, directly and generally applicable index and we name it ‘epidemic evaluation index’ (EEI), which is constructed by 7-day moving average of the log-transformed daily new cases (LMA). EEI could be used to support the decision-making process and epidemic prevention and control strategies through the evaluation of the current epidemic situation. First, we used SARS epidemic data from Hong Kong in 2003 to verify the practicability of the new index, which shows that the index is acceptable. The EEI was then applied to the COVID-19 epidemic situation analysis. We found that the trend direction of different districts in China changed on different date during the epidemic. At the national level and at local Hubei Province level, the epidemic both peaked on February 9. While the peak occurred relatively earlier, i.e. on February 5 in other provinces. It demonstrated the effectiveness of decisive action and implementations of control measures made by Chinese governments. While local governments should adjust management measures based on local epidemic situation. Although the epidemic has eased since late February, continued efforts in epidemic control are still required to prevent transmission of imported cases in China. However, the global COVID-19 epidemic outside China continues to expand as indicated by the EEI we proposed. Currently, efforts have been made worldwide to combat the novel coronavirus pandemic. People all over the world should work together and governments of all countries should take efficient measures in the light of Chinas experience and according to national circumstances and local conditions.

## 1 Introduction

In late December 2019, an outbreak of pneumonia occurred in Wuhan, China, and subsequently spread throughout the country. Soon, the pathogenic virus was proven to be a novel coronavirus (COVID-19), which shares 79.5% sequence identity to SARS-CoV(Zhou et al. 2020a). A total of 81,498 laboratory-confirmed cases, including 3,267 deaths (4.0%) had been reported in China by March 22, 2020; meanwhile, 210,644 laboratory-confirmed cases and 9,517 deaths (4.5%) were reported outside China(WHO 2020). Common symptoms of COVID-19 infection include fever, fatigue and dry cough. Some patients would experience severe complications, including acute respiratory distress syndrome (ARDS), arrhythmia and shock(Wang et al. 2020). According to a research, the mean incubation period is 3.0 days(Guan et al. 2020). Obviously, the COVID-19 epidemic can spread rapidly by human-to-human transmission, but the exact epidemiological characteristics and the specific modes of transmission remain partially unknown. The basic reproduction number (R0) was used to evaluate the transmission ability of COVID-19. Riou et al estimated that the R0 of COVID-19 in January was about 2.2, suggesting the possibility for continuous human-to-human transmission(Riou and Althaus 2020). Zhou et al obtained similar results with R0 between 2.8 and 3.3(Zhou et al. 2020b). Lipsitch et al. reported that Severe Acute Respiratory Syndrome-associated Coronavirus (SARS-CoV) had an R0 value of approximately 2.2 to 3.6(Lipsitch et al. 2003). The transmission capacity of COVID-19 appeared to be similar or higher compared to SARS-CoV. As a continuous unconventional public health event that can lead to economic recession and health damage, it is of great importance to timely estimate the epidemic stage of the outbreak and determine whether it is possible to return to normal work and life. Faced with such a sudden outbreak of new infectious diseases, we have no history to learn from and no evidence to count on. The traditional epidemic prediction models, including SEIR, are highly sensitive to the pre-specified epidemiological parameters, and the prediction results may be highly inconsistent due to different assumptions and data sources. The reliability of the SEIR model can be affected by many factors that are difficult to estimate, including seasonality of coronavirus, inter-city mobility, transmission of travel behavior, and the number of asymptomatic infections. The model may underestimate the scale of the outbreak(Wu et al. 2020). Traditional models prove to be most reliable only under ideal conditions.

To eliminate these limitations and build a practical index based on public reported data to timely evaluate current epidemic situation, we used moving average of log-transformed daily new cases (LMA) to establish a specific index named ‘epidemic evaluation index’ (EEI). The EEI was verified through previous SARS epidemic data in 2003 and then adopted to evaluate the present COVID-19 epidemic.

## 2 Methods

We obtained the epidemic data in China available from January 16 to March 22, 2020 in Chinese public domain(CCDC 2020; NHC 2020) and the data outside China from January 27 to March 22, 2020 on WHO website(WHO 2020). Based on public reported data, we calculated two parameters to evaluate the epidemic situation and demonstrate the epidemic trends.

### 2.1 LMA

The pneumonia caused by COVID-19 has a median incubation period of 3.0 days(Guan et al. 2020). The median time from the onset of symptom to hospital admission was 7.0 days(Wang et al. 2020), and it should become shorter due to the deeper public awareness of the disease. According to NHC, the average time from onset to diagnosis is 4.95 days by February 17, 2020, so the number of new cases per day cannot accurately account for the epidemic situation. The choice of LMA duration can be changed based on the dynamics of the epidemic. Here, we mainly used the 7-day LMA, which can better cover both the incubation period and the duration from the first symptoms to diagnosis. Other MAs (5-day, 10-day and 14-day) were used for sensitivity analysis and auxiliary display was performed in the appendix.

Because the inclusion criteria for the daily new confirmed cases in Hubei Province has changed for several times, we firstly added 1 to the actual observation, and then performed a natural logarithmic transformation to make the index more tolerant of outliers. Moreover, the index after data transformation has better stability. After data transformation, we get the natural logarithm of actual observation as LN_j_:

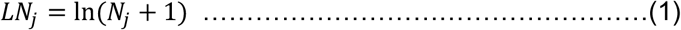

Thus, the LMA was read as average of LN_j_:

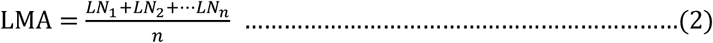

*N*_*1*_, *N*_*2*_,*…, N*_*n*_ indicates the daily new cases for the past n days (e.g. 7-day LMA on February 7 = (Sum of log-transformed daily new cases on February 1 to February 7) /7)

When analyzing epidemic situation, actual observation was presented in logarithmic transformation form and LMA curve was shown in the figures.

### 2.2 Epidemic evaluation index (EEI)

We have predicted the COVID-19 epidemic trend in different districts of China based on moving average and its prediction limits(He et al. 2020). Furthermore, we constructed a new index to more intuitively reflect epidemic situation.

Taking a 7-day interval as an example, in this scenario LMA1 indicates 7-day LMA on a certain day (*X1*: *LN*_*1*_ to *LN*_*7*_), and LMA0 indicates LMA for the previous day (*X0: LN*_*0*_ *to LN*_*6*_).

We defined an ‘epidemic evaluation index’ (EEI) to evaluate epidemic situation: mean and variance of the EEI are calculated on the basis of Taylor expansion(Seltman)

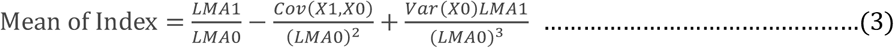

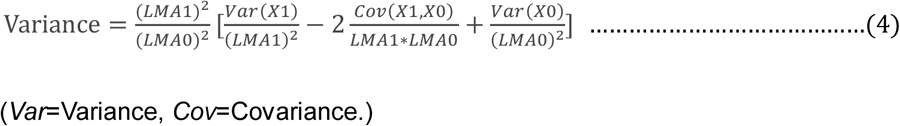

Then, the upper and lower limits of the EEI are calculated by the following formula:

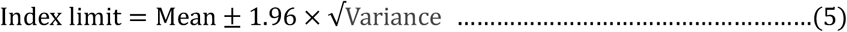

A line chart of the EEI over date can indicate the epidemic situation. The upper and lower limit represent the worst and the best estimates of the epidemic situation, respectively. If the EEI is higher than 1.0, it suggests that the epidemic is still developing. High level restriction measures should be maintained. If the EEI drops below 1.0, it can be judged that the epidemic has entered the period of decline. It’s time to plan to phase out some stringent preventive and control measures. If the EEI remains below 1.0 for more than 7 days (one time interval of LMA), it indicates that the epidemic has entered a stable remission period. It is possible to phase out highly stringent restrictions and then resume production gradually.. If the upper limit of the EEI is less than 1, indicating that under the worst circumstance, the EEI is also at a low level. It can be said with greater confidence that the epidemic will soon be completely eliminated. If there are very few daily new confirmed cases (<5), the EEI will become not applicable. In this scenario, we should evaluate the epidemic situation based on suspected cases or other indicators.

### 2.3 The verification of ‘EEI’

SARS epidemic data of Hong Kong in 2003 was used to verify the practicability of the EEI. The mean incubation period of SARS was 5 to 7 days(Tan et al. 2005), and the average duration of onset of symptoms to hospital admission was 3.8 days(Feng et al. 2009), so we used 10-day LMA and the corresponding ratio to verify that they were consistent with the actual epidemic trend.

## 3 Results

### 3.1 The verification of the new index

In our research, we adopted a new parameter ‘the EEI’ to evaluate the COVID-19 epidemic situation. the EEI is easier to understand than the traditional R0, but it hasn’t been testified. To verify the practicability of the new index, we obtained the epidemiological data in Hong Kong Special Administrative Region from 17 March to 21 May, 2003 available on WHO website(WHO 2003) and from the appendix of a Chinese study(Xu 2019).

The blue curve which represents the change in log-transformed value of actual observation is highly undulating, making it difficult to evaluate the trend. the EEI is more stable and intuitive (figure 1A). The epidemic situation can be judged by directly comparing the index with 1.0. Since May 11 the daily new confirmed cases had dropped below 5, so the EEI was no longer applicable. Based on the number itself, it can be determined that the epidemic was in its final phase. The fluctuations at the end of the epidemic is negligible at this time. Since **April 18**, the the EEI had fallen below 1.0, suggesting the epidemic had reached the peak and began to decline. We can see that from the same day (**April 18**) to the end, the overall SARS epidemic trend had been indeed declining, which verified the conclusion indicated by the EEI.

**Figure 1.**
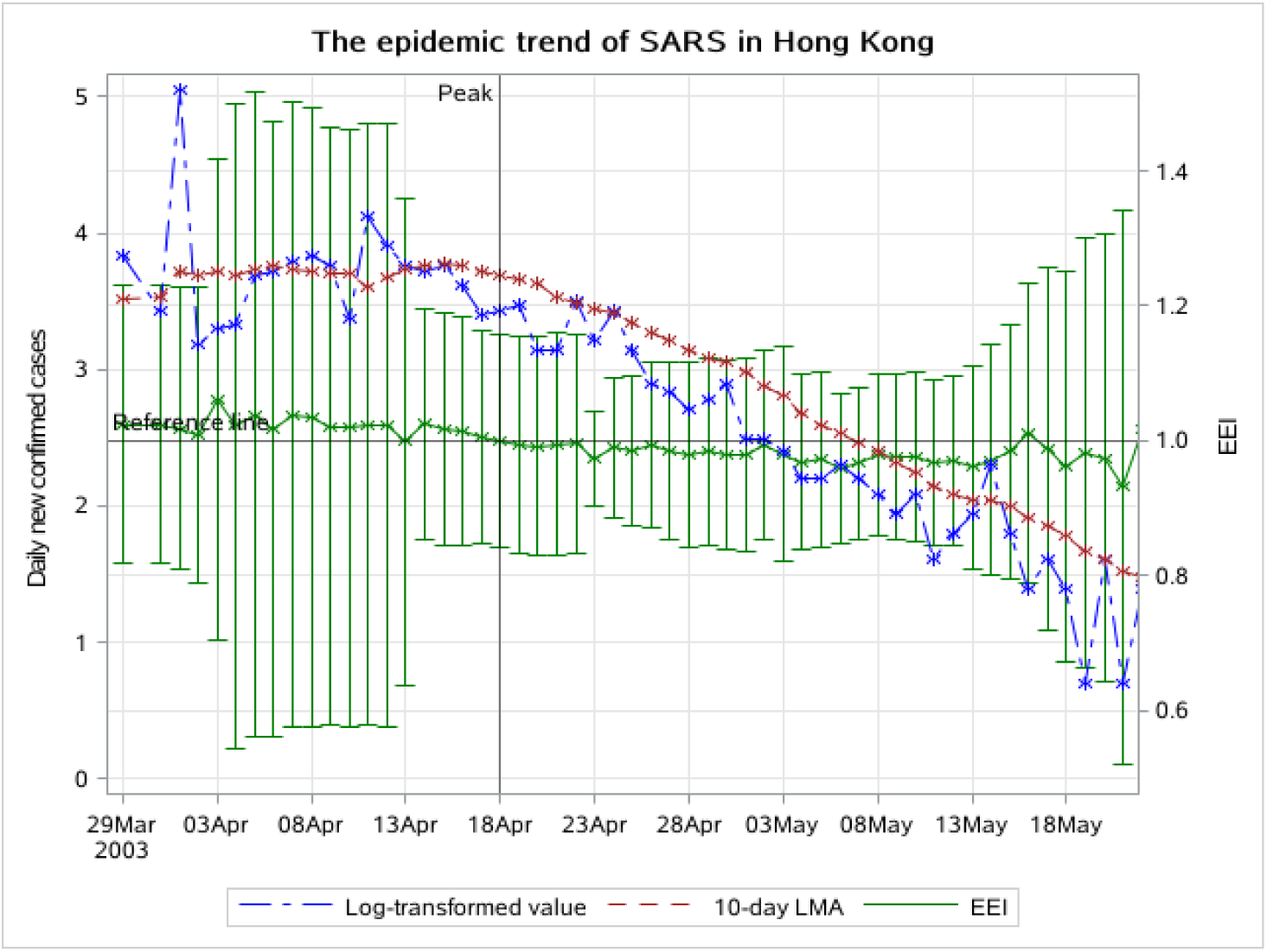
A: The log-transformed value of daily new confirmed cases, 10-day LMA and the EEI in Hong Kong, from Mar 29 to May 21, 2003.

In Hong Kong, SARS began to erupt and the number of cases increased sharply approximately on March 29(Tsang and Lam 2003). Since March 26, the local government had required close contact cases to go to designated hospitals for examination, and since March 29, all schools had been closed for 2 weeks. About 3 weeks (three times the incubation period) later namely on **April 18**, the EEI began to decline. The peak of the epidemic coincided with the actual measures of epidemic prevention and control.

According to previous estimates of the SARS epidemic peak in Hong Kong, different studies have yielded different results. Peiris suggested that on April 25, the epidemic in Hong Kong showed signs of peaking(Enserink 2013). While Lee concluded that the epidemic peaked at the end of March(Lee 2003) and Xu estimated that the epidemic would peak 14 days (on April 14) after the start date (on March 31). It can be seen that the conclusions of previous prediction models are quite different, and it’s difficult to determine which one has a higher degree of reliability when it is applied to epidemic prediction. But now we can see that the epidemic peaked on April 14 is the hypothesis closest to the actual situation, and also the hypothesis closest to our conclusion.

The result indicated by the EEI is verified by the overall SARS epidemic situation, and is consistent with actual progress of measures for epidemic prevention and control. These two aspects show that the reliability of the EEI is the same as the above-mentioned peak prediction method, and is consistent with the actual situation. The index we proposed seems realistic and acceptable.

### 3.2 Evaluation of the COVID-19 epidemic situations

Due to different trends in different districts, we evaluated the COVID-19 epidemic situations at four levels: China, Hubei Province, provinces outside Hubei, and areas outside China. Daily new cases were mainly calculated in laboratory-confirmed category. If there are very few daily new confirmed cases (<5), suspected cases category will be also considered. The epidemic broke out on different dates in different districts, so the start date in the following figures are different for each region.

#### 3.2.1 China

The change in the log-transformed value indicates that it has fallen since February 4 (figure 2A). But it continued to fluctuate for several days, which made trends difficult to evaluate. Since February 7, the increase in 7-day LMA has slowed significantly, peaked on February 9, and then began to decline. The EEI has been less than 1 since February 10. We can see that the index increased slightly on February 18 and February 22, in part because the NHC data included not only laboratory-confirmed cases, but also clinically diagnosed cases over several days. Since the number of new cases in each category was not stated, they cannot be distinguished. The index remained below 1.0 until February 27. On February 28, the index rose to slightly above 1.0, but then soon fell back. From these three aspects, we conclude that the epidemic peak in China was about February 9. The epidemic has entered a period of decline since February 9, and has entered a stable remission period since February 17. On March 16, the EEI rose above 1.0 and didn’t fell back, indicating that a new epidemic period caused by imported cases has begun.

**Figure 2A:**
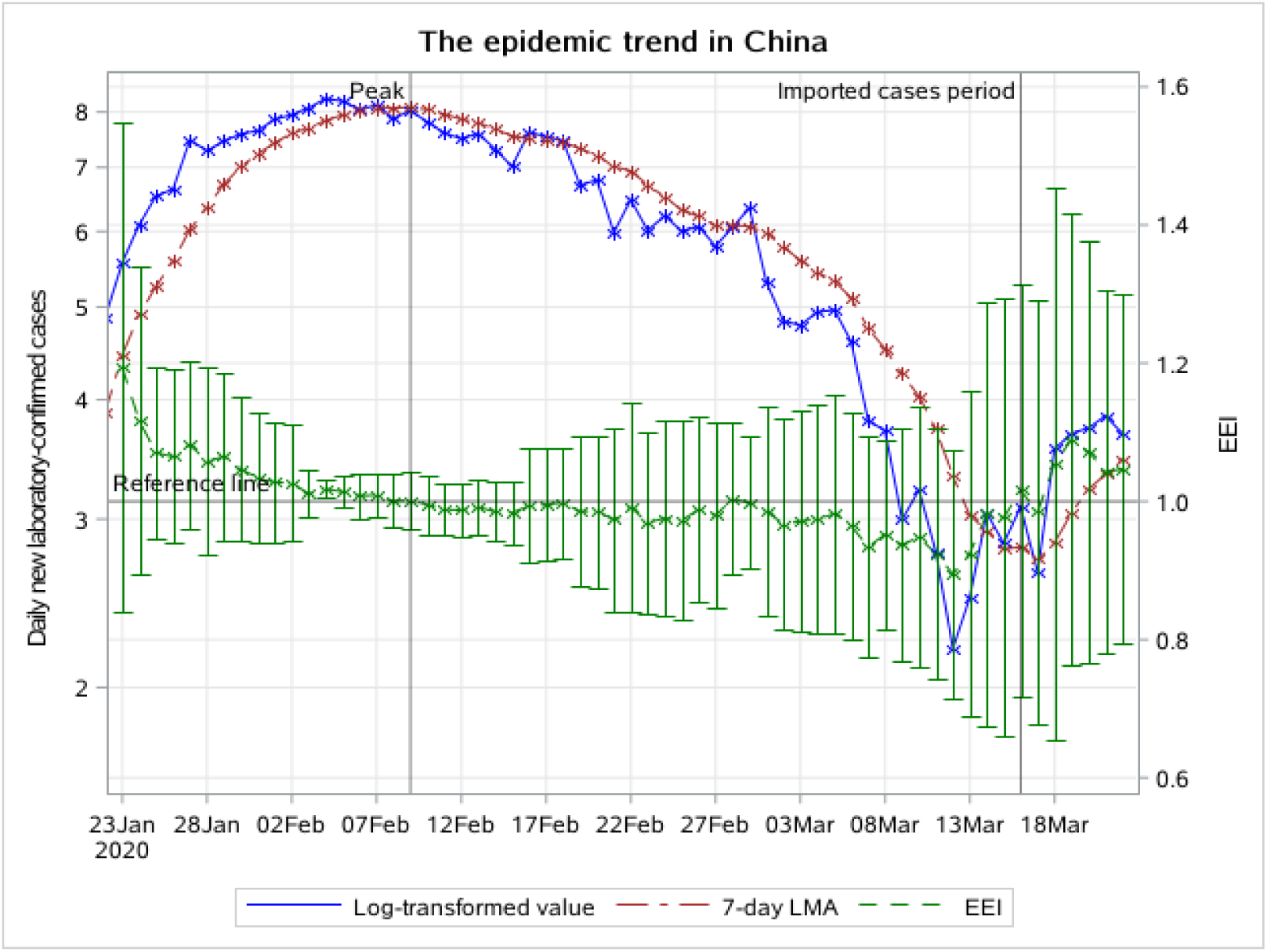
The epidemic trend of daily new laboratory-confirmed cases in China, from January 23 to March 22 2020. The blue line indicates the change of the log-transformed value over date. The brown line indicates the change of 7-day LMA over date. The green line indicates the change of the EEI over date.

### 3.2.2 Hubei Province

Because Hubei Province accounts for the majority of cases, the epidemic in Hubei Province is similar to that in the country. On February 9, both the log-transformed value of actual observation and 7-day LMA started to decline, which is the same as the whole country (figure 3A). Since February 10, the EEI of laboratory-confirmed cases has been less than 1.0. We can see that the index was slightly higher than 1.0 on February 18 and February 28, because NHC data included not only laboratory-confirmed cases but also clinically diagnosed cases during days before and after. We consider it a normal fluctuation. If public health measures prove effective, the epidemic will not usher in a second peak. The peak in Hubei Province was approximately on February 9. The epidemic has entered a period of decline since February 9, and has entered a stable remission period since February 17. The the EEI has not rebounded until the current date (ie. March 22), indicating that there is no epidemic of imported cases in Hubei Province.

**Figure 3A:**
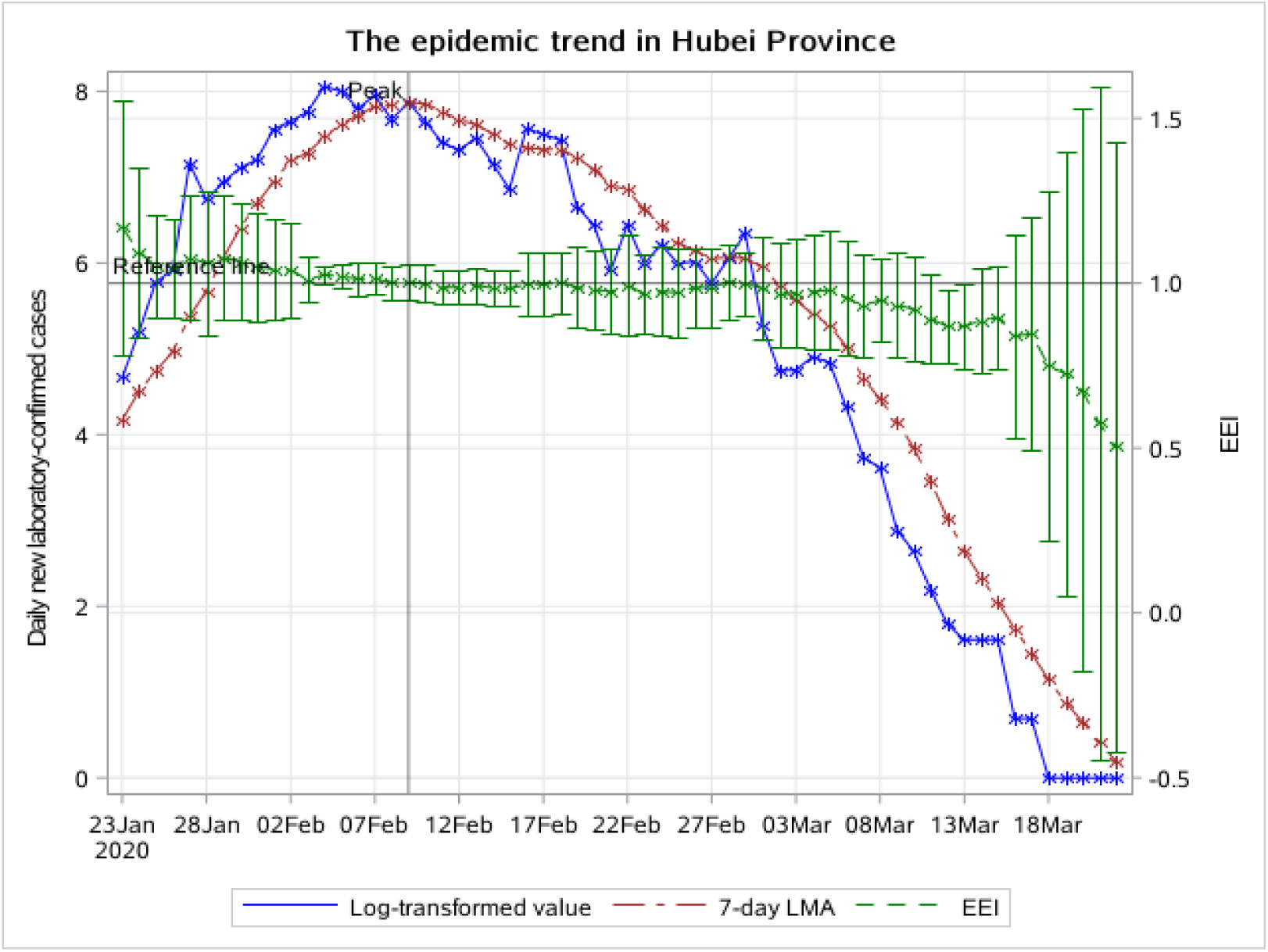
The epidemic trend of daily new laboratory-confirmed cases in Hubei Province, from January 23 to March 14 2020. The blue line indicates the change of the log-transformed value over date. The brown line indicates the change of 7-day LMA over date. The green line indicates the change of the EEI over date.

### 3.2.3 Provinces outside Hubei

The log-transformed value of actual observation peaked on February 3. The 7-day LMA has fallen since February 5 in provinces outside Hubei (figure 4A). The peak appeared earlier than Hubei Province. the EEI has been declining since February 4 and was below 1.0 on February 6. In provinces outside Hubei, the peak was approximately on February 5. The index kept going down for a long time. The epidemic has entered a period of decline since February 5, and has entered a stable remission period since February 13. It is worth noting that the upper limit of the index fell below 1.0 on February 13. However, after a period of decline, the index suddenly rose to over 1.0 again because of the massive growth of daily new laboratory-confirmed cases on February 20 and on February 26. The rise on Feb 20 was caused by cluster epidemic in prisons. On February 28, the number of daily new laboratory-confirmed cases dropped to less than 5, so we replaced the EEI of laboratory-confirmed cases with the EEI of suspected cases to evaluate the epidemic situation after February 28.

**Figure 4A:**
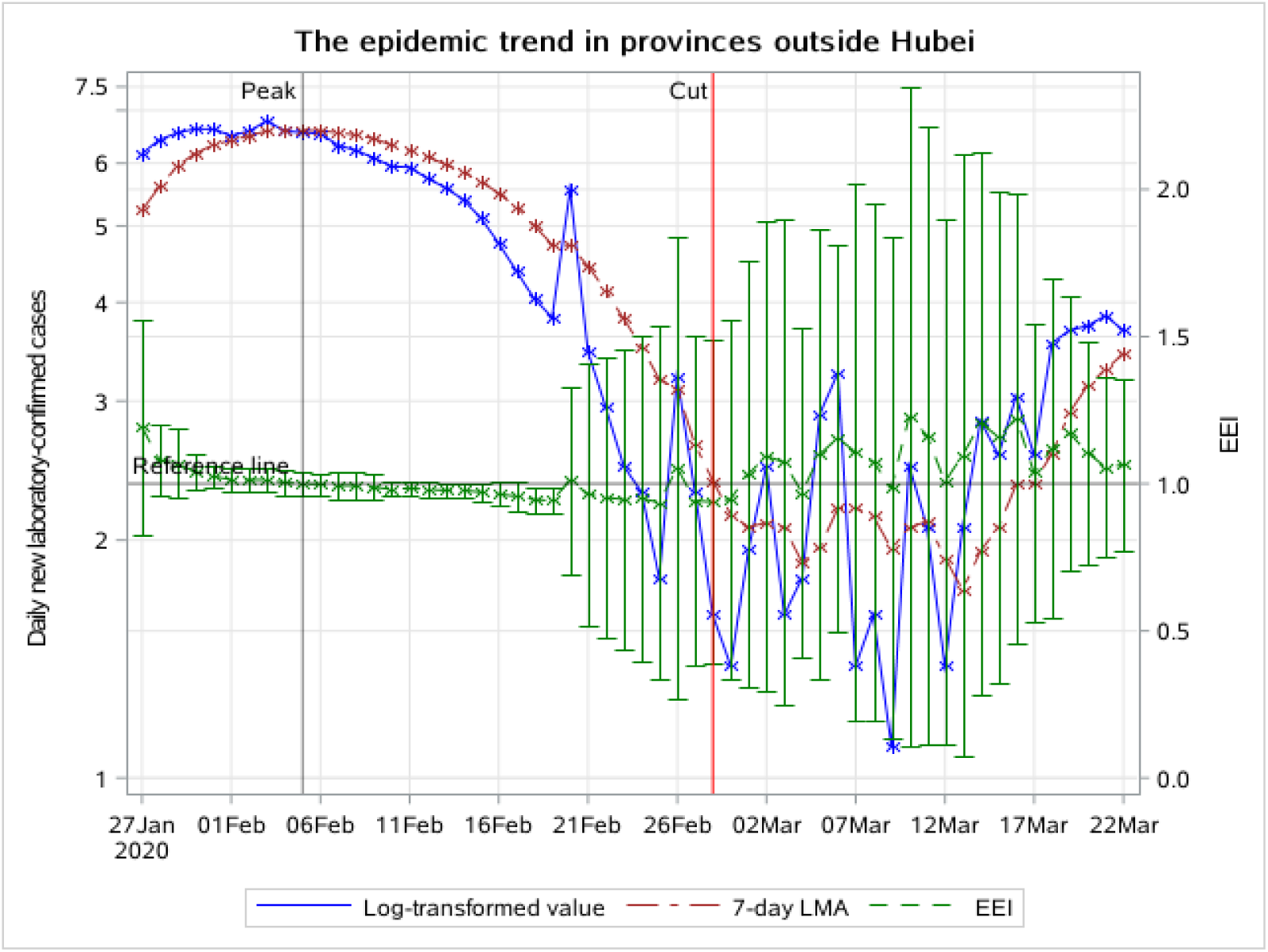
The epidemic trend of daily new laboratory-confirmed cases in provinces outside Hubei, from January 27 to March 22 2020. The blue line indicates the change of the log-transformed value over date. The brown line indicates the change of 7-day LMA over date. The green line indicates the change of the EEI over date.

**Figure 4B:**
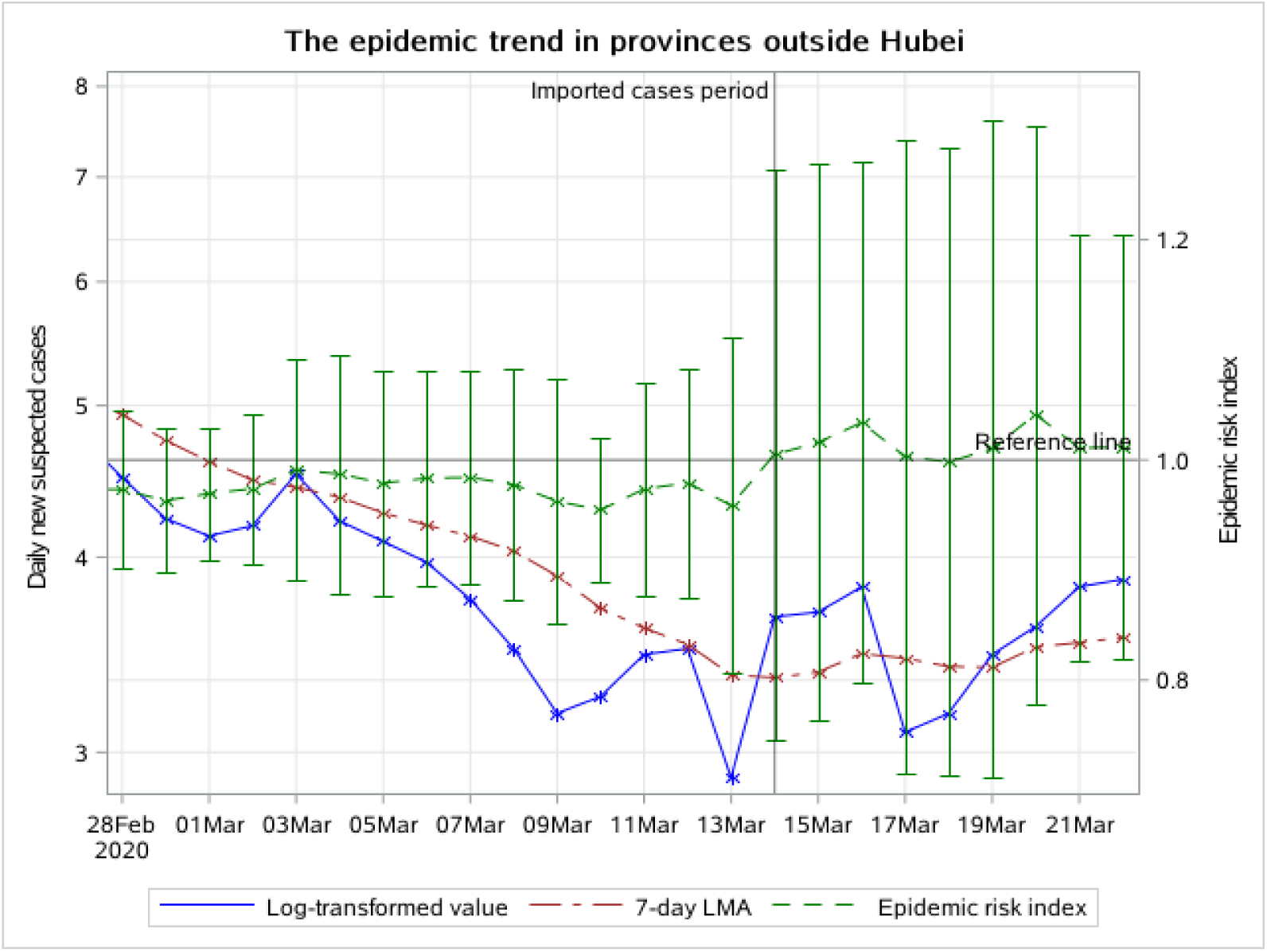
The epidemic trend of daily new suspected cases in provinces outside Hubei, from February 28 to March 22 2020. The blue line indicates the change of the log-transformed value over date. The brown line indicates the change of 7-day LMA over date. The green line indicates the change of the EEI over date.

On March 14, the EEI rose above 1.0, which indicated that a new epidemic period caused by imported cases has begun. The start date of this period is earlier than the whole nation.

### 3.2.4 Areas outside China

Figure 5A shows that the 7-day LMA curve continues to rise, and the EEI is still higher than 1.0 by the current date (ie March 22, 2020). It can be inferred that epidemics outside China are still developing and becoming more serious. Here, we take Republic of Korea, Italy and Iran as examples to evaluate the epidemic situation.

**Figure 5A:**
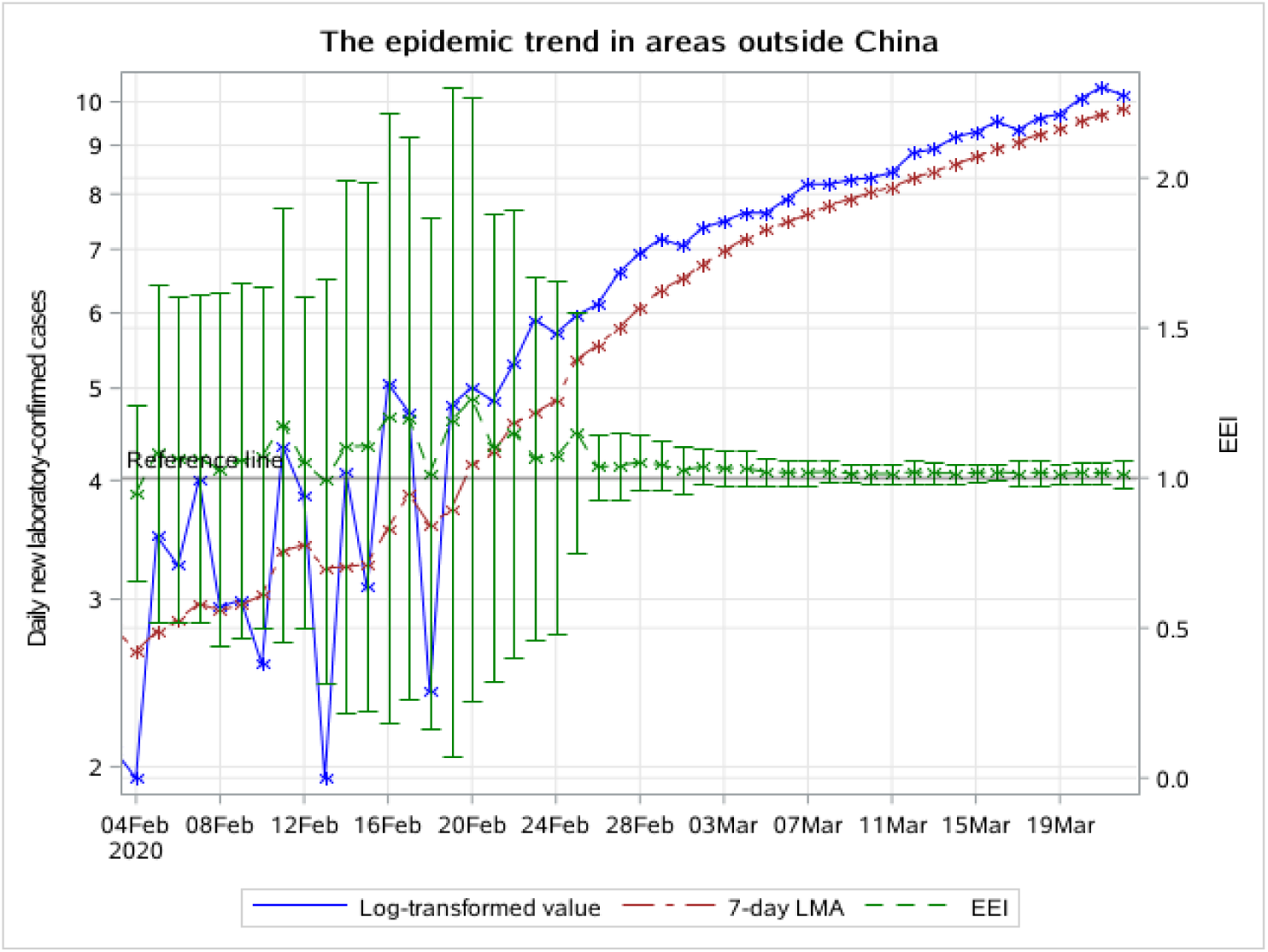
The epidemic trend of daily new laboratory-confirmed cases in areas outside China, from February 4 to March 22 2020. The blue line indicates the change of the log-transformed value over date. The brown line indicates the change of 7-day LMA over date. The green line indicates the change of the EEI over date.

In Republic of Korea, there were very few daily new cases and the EEI was not applicable at the beginning. After February 18, the number of daily new confirmed cases has increased significantly and the EEI has remained above 1.0 until March 4 (figure 5B). At the same time, the 7-day LMA of daily new cases began to decline. We concluded that the peak in Republic of Korea was on March 4. The epidemic has entered a period of decline since March 4, and has entered a stable remission period since March 12. On March 20, the EEI rose above 1.0, maybe due to imported cases.

**Figure 5B:**
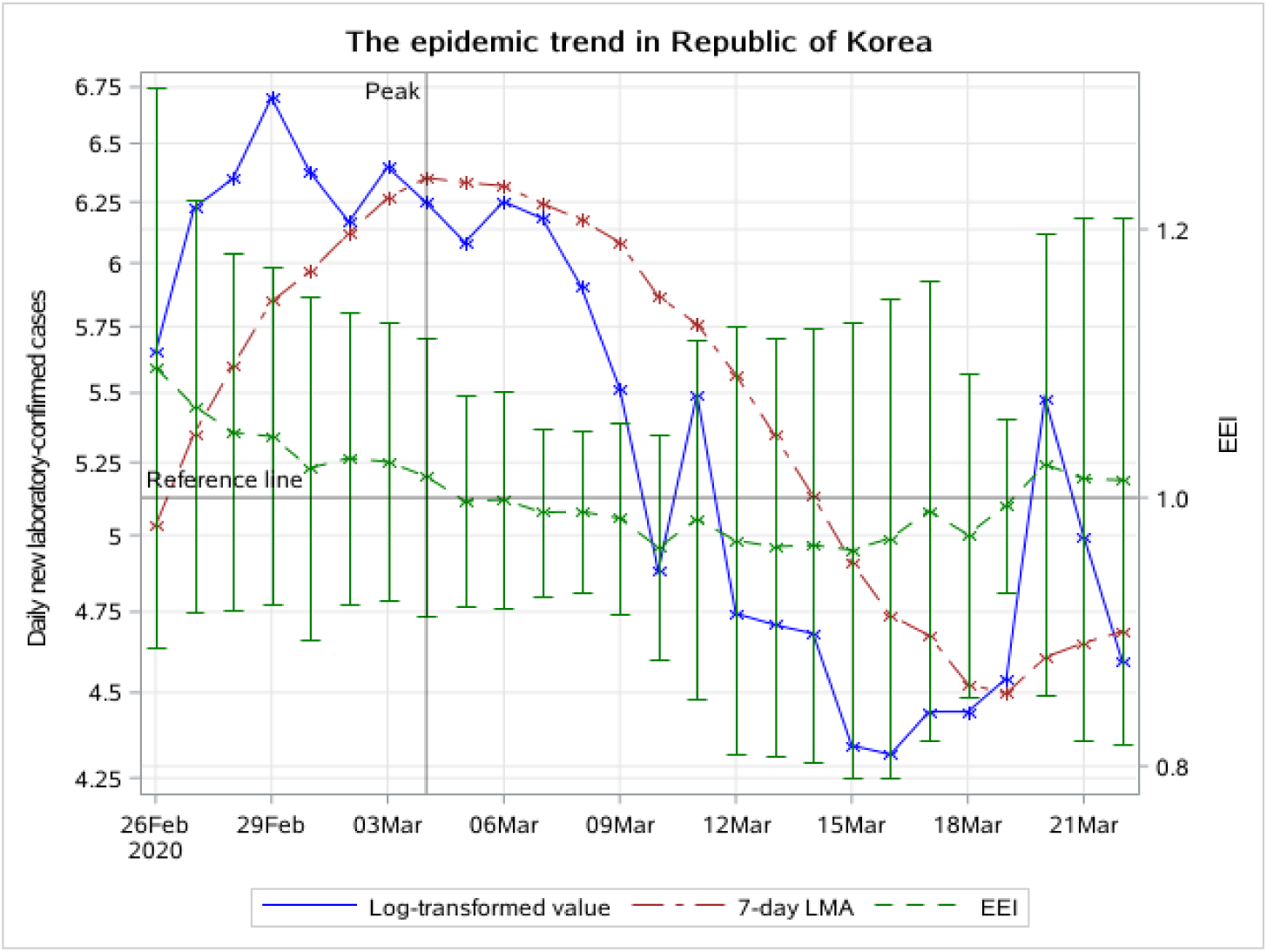
The epidemic trend of daily new laboratory-confirmed cases in Republic of Korea, from February 26 to March 22 2020. The blue line indicates the change of the log-transformed value over date. The brown line indicates the change of 7-day LMA over date. The green line indicates the change of the EEI over date.

In Italy, the EEI is still higher than 1.0, indicating that the epidemic is still developing and hasn’t yet reached the peak (figure 5C).

**Figure 5C:**
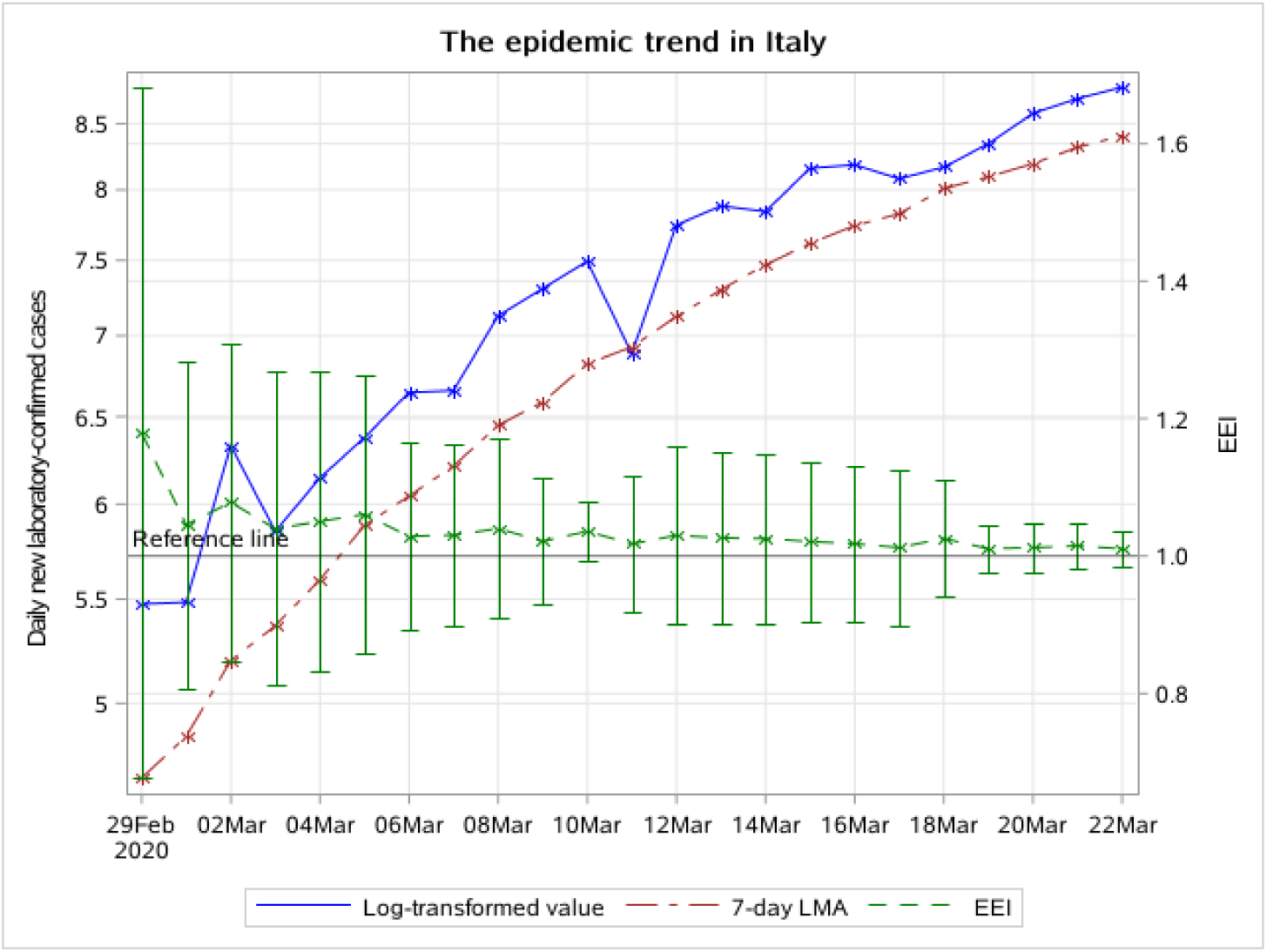
The epidemic trend of daily new laboratory-confirmed cases in Italy, from February 29 to March 22 2020. The blue line indicates the change of the log-transformed value over date. The brown line indicates the change of 7-day LMA over date. The green line indicates the change of the EEI over date.

**Figure 5D:**
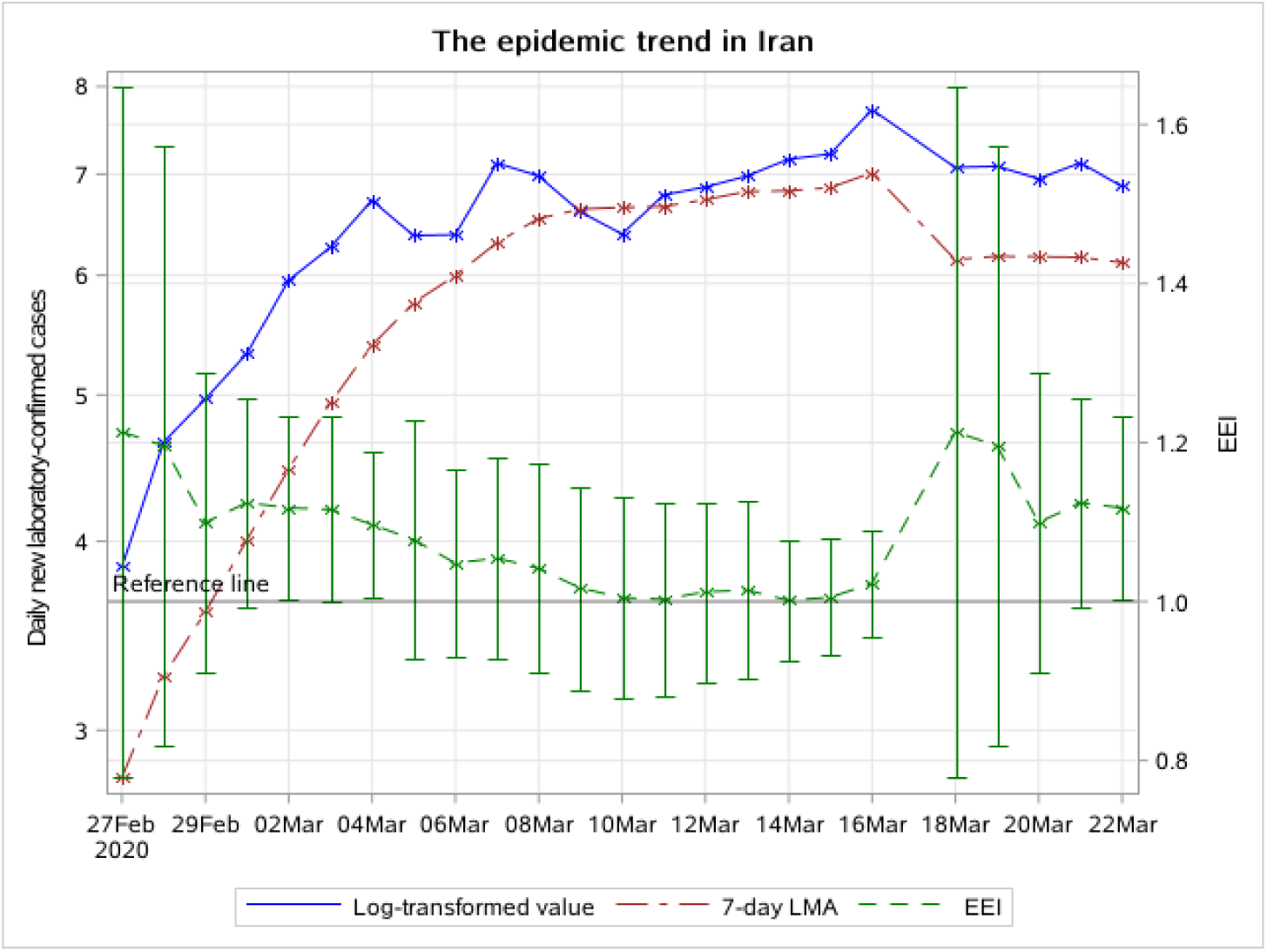
The epidemic trend of daily new laboratory-confirmed cases in Iran, from February 27 to March 22 2020. The blue line indicates the change of the log-transformed value over date. The brown line indicates the change of 7-day LMA over date. The green line indicates the change of the epidemic evaluation index over date

In Iran, the EEI is still above 1.0 until the current date (ie March 22, 2020), which indicates that the epidemic in Iran is still developing. It hasn’t entered the period of decline.

## 4 Discussion and Conclusion

Compared with the EEI curve, the actual observation curve always fluctuates greatly and makes it difficult to judge the trend. The decline in the log-transformed actual observation curve doesn’t mean that the peak has occurred, as it may rise again. The EEI can more accurately reflect the true situation of the epidemic. It aims to comprehensively analyze the epidemic situation based on combination of actual observation and LMA.

Traditional models, including SEIR and Auto Regressive Integrated Moving Average (ARIMA) models, require pre-set application conditions. As mentioned above, the reliability of SEIR model is affected by the seasonality of coronavirus, population mobility, and asymptomatic infection status. The ARIMA models can only be used when the data is stable. The new EEI we proposed avoids these limitations and is more intuitive and reliable with only officially released data. The EEI aims to monitor and evaluate the epidemic in time, not to make predictions. It adopts another idea to provide timely decision support for epidemic prevention strategies.

Regarding the COVID-19 epidemic situation, it can be concluded that whether in Hubei Province or throughout the country, the trend has entered a period of decline due to many measures taken in time. In 2003, public health measures including individual isolation, quarantine, social distancing and community containment once played decisive roles in controlling the SARS epidemic(Wilder-Smith and Freedman 2020). During the COVID-19 epidemic, the Chinese government learned lessons from the experience of SARS epidemic and adopted emergency measures related to population prevention and clinical treatment after the outbreak. Compared with the prevention and control of previous SARS epidemic, significant progress has been made during the COVID-19 epidemic (Guan and Xian 2020). On January 20, 43 days after the first unexplained pneumonia case occurred, traffic control was implemented in Wuhan. 3 days later, on January 23, Wuhan city was closed. 2 days later, on January 25, all provinces in China except Tibet triggered a level 1 public health emergency response. As can be seen from figure 2A, 2B, 3A and 3B, the peak of daily new laboratory-confirmed cases in mainland China and Hubei province were on February 9, approximately 14 days (one quarantine observation period) after the public health emergency response, which indicates that the public health measures proved to be effective. The peak in other provinces appeared earlier, approximately on February 5. The possible reason is that the epidemic outside Hubei Province is less serious and easier to control. We can see that the first-grade prevention policy (on January 20) hadn’t been carried out until 20 days after the outbreak (on Dec 31). After the SARS epidemic in 2003, Chinas public health system has received attention, but problems that emerged during the initial stage of the COVID-19 epidemic indicated that it still lacks the ability to respond quickly to unexpected infectious diseases and public health emergencies. There is still much room for improvement in the prevention and control system for major public health emergencies (Ding et al. 2020).

The EEI of daily new laboratory-confirmed cases in China has fallen below 1.0. In order to avoid excessive economic and social burden caused by the epidemic restriction measures, localities should take actions according to current epidemic conditions. In Hubei Province, as the initial district of the epidemic outbreak, the EEI rose to slightly more than 1.0 twice. It suggests that after the epidemic peak, Hubei Province had to go through a long transition period before entering the final end phase. The local government can plan to restore production of some enterprises, and plan to normalize residents’ travel and public transportation after the peak (February 9). After February 17, the government can reduce restriction level and gradually restore urban traffic. In provinces outside Hubei, the EEI of daily new laboratory-confirmed cases experienced two brief increases, one of which was caused by the prison epidemic. The upper limit of the index had been below 1.0 for several consecutive days. The EEI of daily new suspected cases has been below 1.0 since the peak. It indicates that the speed of the epidemic to the end in areas outside Hubei was faster than in Hubei Province. However, resumption of work and return trips may lead to an increase in new cases. For example, on February 26 the index temporarily rose above 1.0. Local governments in these districts can gradually withdraw highly tight restrictions and public health emergency response based on low-risk level of epidemic conditions after February 13. At the same time, public places and enterprises should continue to monitor body temperature. If public places like department stores and restaurants are to be opened, disinfection measures should be emphasized. However, on March 14, a new epidemic period caused by imported cases began. Due to the development of epidemics worldwide, the direction of domestic epidemic prevention must be changed to prevent the spread of imported cases.

Outside China, the EEI never dropped below 1.0 from the beginning to the current date (ie March 22, 2020), indicating that the global epidemic is still developing. Countries with a rapidly increasing number of confirmed cases, such as Iran, Republic of Korea and Italy, can learn from Chinas prevention and control experience and integrate it with their own economic situation and socio-cultural background. Governments in these countries should value public health measures, such as closing public places, restricting urban traffic and delaying school.

Republic of Korea’s measures to control the epidemic have initially shown effectiveness according to the present EEI. On February 23, Republic of Korea raised the level of COVID-19 epidemic warning to the highest level. Ten days later, on March 4, the epidemic reached its peak. However, there is still a need to monitor the subsequent development of the epidemic. In Italy, the epidemic is still on the rise. Government has shut down the entire country on March 10. If strictly enforced, it can be expected that the epidemic in Italy will become moderate. In Iran, the EEI dropped almost below 1.0 on Mar 14, but it rose again on Mar 18. Further efforts are needed to eventually control the epidemic.

In conclusion, it can be inferred that in China, the peak of the phase-one epidemic has already occurred. However, disinfection, isolation, personnel registration and other effective measures must not be stopped. The practice of China in preventing and controlling the COVID-19 epidemic has offered an important lesson to other countries. In areas outside China, many countries are still facing severe challenges in controlling the epidemic. While the present epidemic situation in Republic of Korea indicates that the epidemic can be contained if efficient prevention and control measures are taken as soon as possible. Although the epidemic has shown significant relief in some districts, as long as the epidemic is uncontrolled anywhere in the world, we cannot fully return to normal life.

Currently, in addition to the challenge for epidemic control, another crucial issue is to admit and treat critically ill patients as soon as possible. In the early stage of the epidemic, clinical referral system, specific medical treatments and adequate medical supplies can help alleviate the cumulative pressure from the confirmed cases within a short period of time. As Bill Gates puts it, “No one could predict what the chance of a new virus emerging was”(Gates 2020). This is an unprecedented situation. This is a common enemy against humanity at this point. The only way we can defeat this pandemic is through solidarity(Ghebreyesus 2020). All countries in the world should work together through international cooperation and take timely actions to slow and stop the spread of COVID-19 and take control of the novel coronavirus pandemic.

## Data Availability

All data generated or used during the study are available on the official website and an appendix of a public research.

http://2019ncov.chinacdc.cn/2019-nCoV/

http://www.nhc.gov.cn/xcs/xxgzbd/gzbd_index.shtml

https://www.who.int/emergencies/diseases/novel-coronavirus-2019/situation-reports/

## Appendix

Sensitivity analysis was performed using different MAs (5 days, 10 days, and 14 days), and supplementary display is provided in the appendix. When comparing MA curves of different time intervals, MA is presented in its actual value.The 5-day, 7-day, 10-day and 14-day MA of daily new laboratory-confirmed cases are presented as below (figure S1-S4). Comparison of MA curves of different durations shows their respective characteristics.

**Figure S1:**
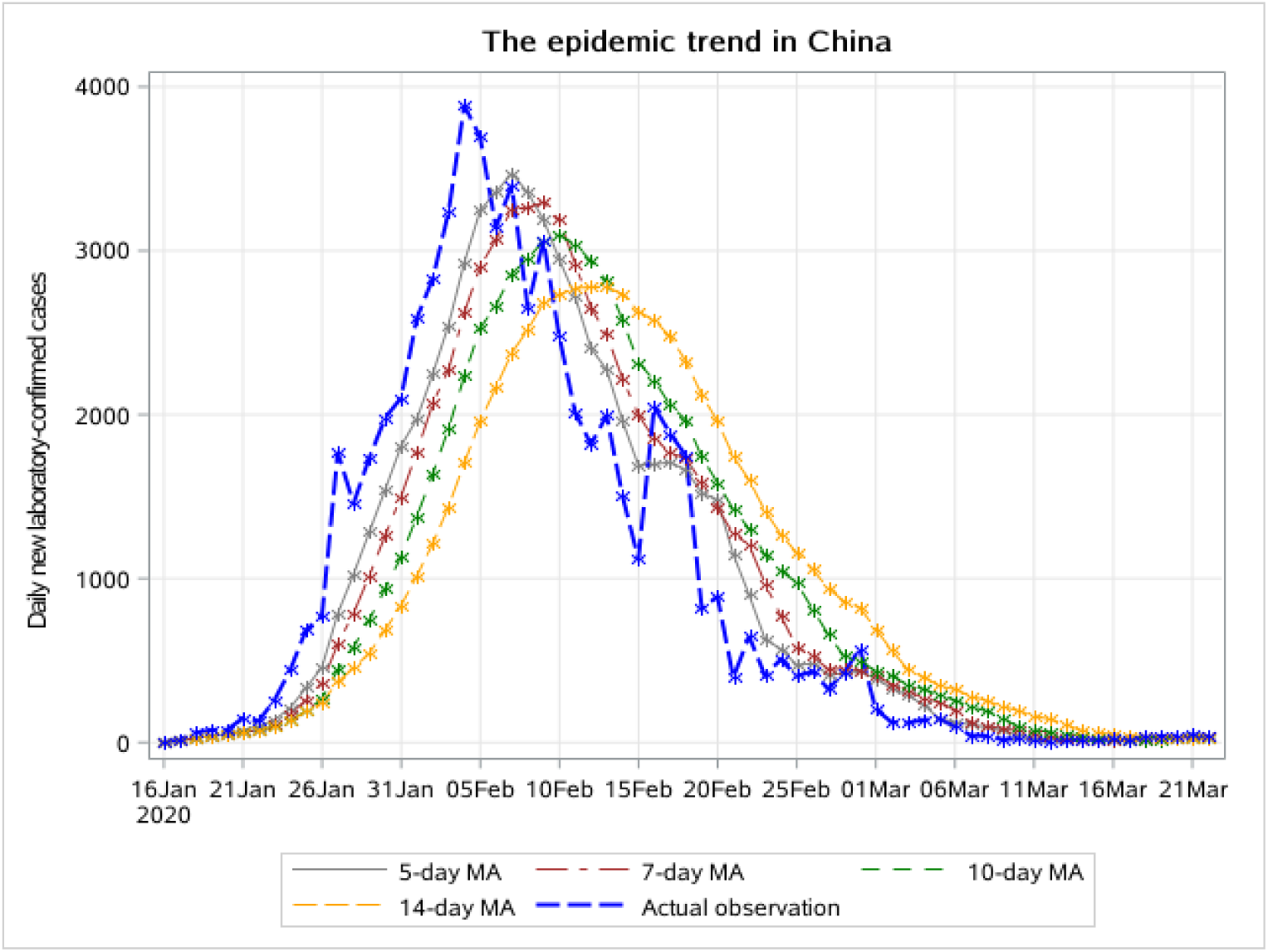
The 5-day, 7-day, 10-day and 14-day MA of daily new laboratory-confirmed cases in China, from January 16 to March 14 2020.

**Figure S2:**
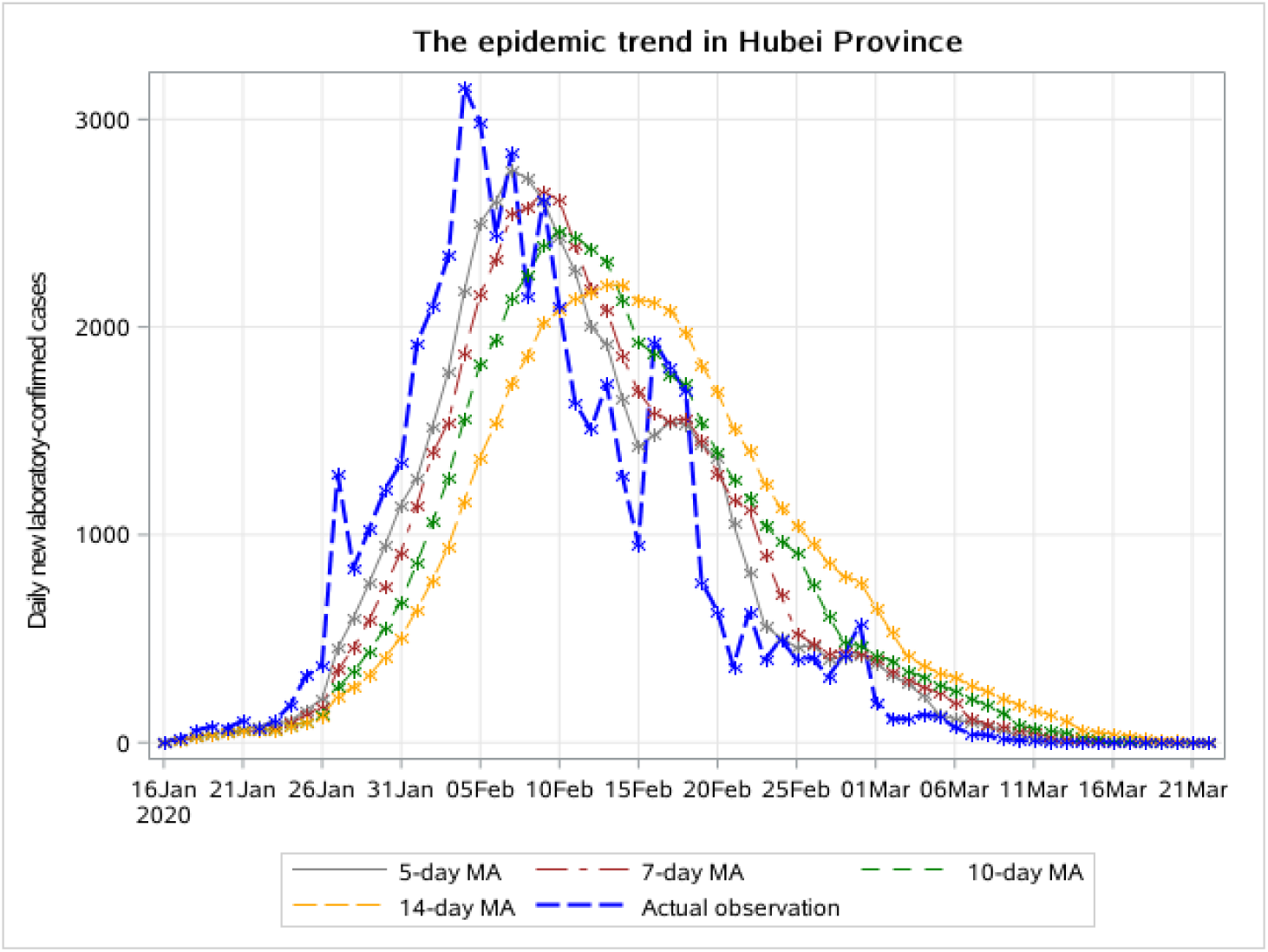
The 5-day, 7-day, 10-day and 14-day MA of daily new laboratory-confirmed cases in Hubei Province, from January 16 to March 14 2020.

**Figure S3:**
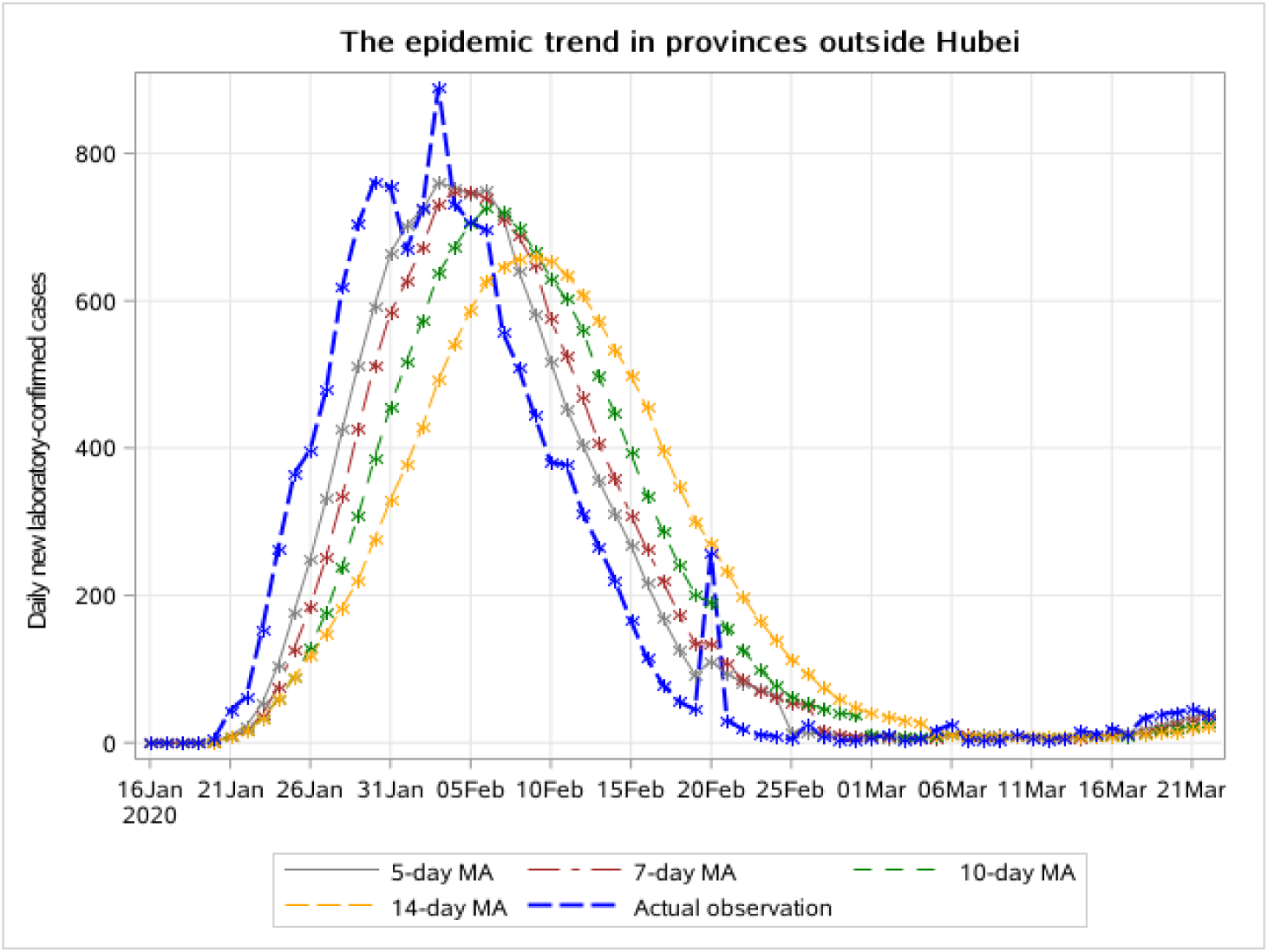
The 5-day, 7-day, 10-day and 14-day MA of daily new laboratory-confirmed cases in provinces outside Hubei, from January 16 to March 14 2020.

**Figure S4:**
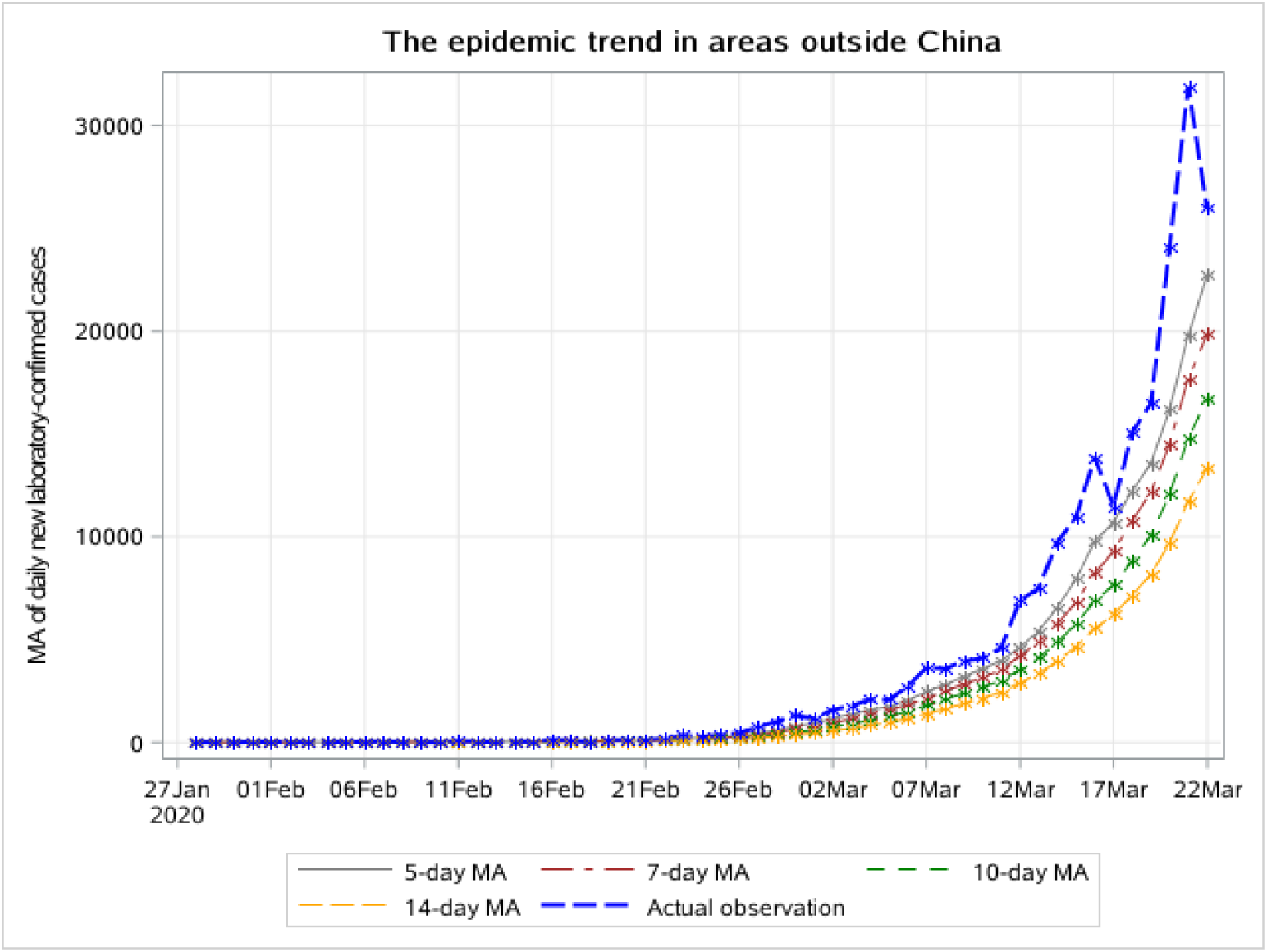
The 5-day, 7-day, 10-day and 14-day MA of daily new laboratory-confirmed cases in areas outside China, from February 4 to March 14 2020.

Figure S1-S3 show that all curves have experienced the turning points. It can be inferred that the epidemic peak in China has passed. Figure S4 indicates that the epidemic outside China is still developing.

Comparison of different MA curves indicates that the longer the interval, the later the peak appeared. The 14-day MA curve appears to be the most stable curve and the 5-day MA curve is the most unstable one. We can use MA of different time intervals to evaluate trends based on the current public health strategy and the urban traffic situation at different stages. Regardless of the time interval of MA, the peak of the epidemic in provinces outside Hubei always appears earlier than that in Hubei Province, which is consistent with our conclusion based on 7-day LMA.

